# Role of non-aerosols activities in the transmission of SARS-Cov-2 infection among health care workers

**DOI:** 10.1101/2021.04.22.21255922

**Authors:** Christophe Paris, Emilie Tadié, Christopher Heslan, Pierre Gary-Bobo, Sitty Oumary, Anaïs Sitruk, Pascal Wild, Pierre Tattevin, Vincent Thibault, Ronan Garlantezec

**Affiliations:** CHU de Rennes, Univ Rennes, Inserm, EHESP, Irset (Institut de recherche en santé, environnement et travail) - UMR_S 1085, F-35000 Rennes, France; CHU de Rennes, F-35000 Rennes, France; INRS, F-54500 Vandoeuvre Lès Nancy, France; CHU de Rennes, Univ Rennes, INSERM U1230, IFR140, F-35033, Rennes, France

**Author notes:** **Corresponding author Corresponding author: Christophe Paris, MD, PhD**, University Hospital Rennes, Occupational Diseases Department – Le Chartier, 2 Rue Henri Le Guilloux, F-35033 Rennes Cedex 09, France.

## Abstract

**Background:** Since the emergence of SARS-CoV-2, health care workers (HCWs) have been on the front line in caring for COVID-19 patients. Better knowledge of risk factors for SARS-CoV-2 infection is crucial for the prevention of disease among this population.

**Methods:** We conducted a seroprevalence survey among HCWs in a French university hospital after the first wave (May-June 2020), based on a validated lateral flow immuno-assay test (LFIAT) for SARS-CoV-2. Demographic characteristics as well as data on the working characteristics of COVID-19 and non-COVID-19 wards and 23 care activities were systematically recorded. The effectiveness of protective equipment was also estimated, based on self-declaration of mask use. SARS-CoV-2 IgG status was modelled by multiple imputations approach, accounting for the performance of the test and data on serum validation ELISA immunoassay.

**Findings:** Among the 3,234 enrolled HCWs, the prevalence of SARS-CoV-2 IgG was 3.8%. Contact with relatives or HCWs who developed COVID-19 were risk factors for SARS-CoV-2 infection, but not contact with COVID-19 patients. In multivariate analyses, suboptimal use of protective equipment during naso-pharyngeal sampling, patient mobilisation, clinical and eye examination was associated with SARS-CoV-2 infection. In addition, patients washing and dressing and aerosol-generating procedures were risk factors for SARS-CoV-2 infection with or without self-declared appropriate use of protective equipment.

**Interpretation:** Main routes of transmission of SARS-CoV-2 IgG among HCWs were i) contact with relatives or HCWs with COVID-19, ii) close or prolonged contact with patients, iii) aerosol-generating procedures.

## INTRODUCTION

Since the emergence of SARS-CoV-2 at the end of 2019 in Wuhan, China, healthcare workers (HCWs) have been on the front lines of the pandemic. Previous publications have reported high percentages of HCWs among patients with coronavirus disease 19 (COVID-19). Initial reports from China found that HCWs represented 3.8% of all cases (1,716 HCWs/44,674) but this proportion was 29% of HCWs in patients admitted for severe pneumonia ^1^. In the USA, HCWs represented 16% of the 315,531 cases of COVID-19 among individuals with a known occupational status^2^. According to the European CDC, the proportion of HCWs among COVID-19 cases varied from 9% to 26% in several EU countries with available data^3^. Finally, a recent meta-analysis reported an overall seroprevalence of SARS-CoV-2 antibodies among HCWs of 8.7% 95CI 6.7-10.9% ^4^. In France, 67,811 HCWs were infected by SARS-CoV-2 between March 2020 and February, 2021^5^. In this context, understanding the main routes of transmission of SARS-CoV-2 in HCWs is an important public health question. Several care activities are known or suspected to be associated with increased risk of transmission of several coronaviruses (SARS, MERS and SARS-CoV-2), in particular aerosol-generating activities such as intubation, high-flow oxygen and mechanical ventilation^6, 7^. In France, a recent study including 2,329 infected HCWs reported that masks were not worn during eye examinations in 47.6% of cases, or during high-risk activities in 19.4% of cases^8^. Other circumstances of possible contamination among these HCWs were the initial absence of recommendations to wear masks in care settings or the use of protective equipment only with suspected or confirmed COVID-19 patients ^9, 10^. Current knowledge on the pathways of SARS-CoV-2 transmission among HCWs remains limited, and requires further analysis. We aimed to precisely assess main care activities associated with the risk of SARS-CoV-2 infection in HCWs by performing a large sero-prevalence study associated with questionnaires on contacts, and practices in a French university hospital, after the first wave.

## METHODS

### Population and data definitions

Between May, 29^th^ and July, 10^th^ 2020, we conducted a sero-epidemiological study at the Rennes University Hospital, a 1,500-bed tertiary care centre in western France, which served as a referral centre for COVID-19 during the first epidemic wave (population catchment area, 1.5 million inhabitants). All HCWs working in the hospital (n=8,540) were invited to be tested for COVID-19, with a finger-prick rapid test (Figure 1). At inclusion and before the realization of the test, they respond to a short questionnaire with data on socio-demographic characteristic (age, sex, occupation, ward) and symptoms and potential risk factors (n=7,003, 82% participation rate). For 1,832 HCWs working in COVID-19 wards and a random sample of HCWs working in non COVID-19 wards (n=1,421), a supplemental questionnaire concerning occupational exposure was addressed. (Figure 1). In this supplemental questionnaire, we collected data about the following activities during working hours: i) patients care (consultation, vital sign measurement, insertion of central or peripheral venous catheters, naso-gastric tubes, and/or urinary catheters, assistance during delivery, loco-regional anaesthesia, clinical examination, naso-pharyngeal sampling, oral, eye, and ear, nose, throat (ENT) examination, aerosol-generating procedures, patient mobilisation, bed making, feeding, surgery, distribution of drugs, washing, dressing, mouth care, mobilisation and respiratory physiotherapy and dental treatment), ii) shared activities with other staff during working hours (transmissions, mealtimes, breaks, and meetings and changing-room habits). The use of protective equipment during these activities was also investigated (masks, gloves, lab coats, etc.). Mask use was categorized as appropriate if the HCWs kept the masks, whatever its type, throughout the activity, sub-optimal if masks were irregularly use, and absent otherwise.

Serological status. After completion of the questionnaire, all HCWs underwent a SARS-CoV-2 Lateral Flow ImmunoAssay Test (LFIAT), namely the NG-Test^®^ finger-prick test^11^, with a reading 20 min after the prick by trained nurses or doctors. This test allows the detection of anti-SARS-CoV-2 IgG with a sensitivity of 82.5% and a specificity of 98% ^12^. If the LFIAT was positive, a venous blood sample (7 mL) was drawn to confirm the serological status (Wantai SARS-CoV-2 Ab ELISA, Beijing, China). Indeed, as the prevalence of SARS-CoV-2 IgG in the Western France during the study period was estimated to be 2%, we expected a low positive predictive value but a good negative predictive value. All data were stored in an on-line database using SPHINX^®^. The study obtained the agreement of the Lyon Institutional Review Board (May, 28^th^ 2020). All HCWs signed an informed consent form, and the study was recorded on ClinicalTrials.gov (#35RC20_9716).

### Statistical Analysis

The clinical COVID-19 status was defined as probable if patients presented with fever, dyspnoea, and at least one of the following: cough, myalgia, headache or unusual fatigue, or if patients presented with anosmia or ageusia. Patients with other symptoms were defined as possible COVID-19.

Our validation study ^12^ demonstrated that SARS-CoV-2 serological test with LFIAT had a predictive positive value of 49.7% and a negative predictive value of 99.7% in our population study for IgG. We thus determined the SARS-CoV-2 serological status (negative / positive) using two distinct approaches. First, only positive IgG LFIA tests were retained to define a positive status. However, as a suitable proportion of HCWs also underwent a Wantai SARS-CoV-2 Ab ELISA as a control, we decided to retain this last result when available, irrespective of the LFIA result. All negative IgG LFIA tests were considered negative. This definition is thereafter referred to as ‘by combination’, and was used in all descriptive and univariate analyses as well as multivariate analyses. Second, we proceeded to multiple imputation (n = 50 databases) of the SARS-CoV-2 serological status using a logistic regression model based on the clinical COVID-19 status (defined above) for subjects with a positive IgG LFIA test (except for subjects with a Wantai SARS-CoV-2 Ab ELISA result). Conversely, negative IgG LFIA tests were randomly imputed positive at a 0.3% frequency, corresponding to the false negative rate with this test in our validation study. This definition is thereafter referred to as ‘by multiple imputation’ and only used when multiple approaches were applied.

An analysis of risk factors associated with the serological status was performed using logistic regression models. The descriptive and primary analyses used the ‘by combination’ definition of serological SARS-CoV-2 status, whereas the analyses of overall and occupational risk factors used both definitions.

Factor analysis was first performed for the analysis of activities, as preliminary analyses demonstrated strong correlations among these data (not shown). Six factors were defined corresponding to nurse, auxiliary-nurse, medical, surgical, physiotherapist and ENT activities. Associations between a positive SARS-CoV-2 status and the use of protective equipment were then analysed by separate backward logistic regression models according to each factor. All models were adjusted for age, sex, and occupation and, depending on analyses, contact with COVID-19 patients or relatives with COVID-19.

Statistical analyse were performed using the SAS^®^ package, v9.4, with FACTOR, LOGISTIC and MIANALYSE procedures. Results are presented as Odds ratio (OR) with their 95% confidence intervals. A p value below 0.05 was considered as significant.

## RESULTS

Among the 3,234 participants that completed the supplemental questionnaire and underwent LFIAT (Figure 1), 120 (3.8%) HCWs presented with IgG SARS-CoV-2 according to the ‘by combination’ definition (Table I). We observed close-to-significant differences (p = 0.06) among occupations, with cleaners, stretcher-bearers and residents having the highest rate of positive tests. There were no significant differences according to sex, age, smoking status, or presence of comorbidities. The number of symptoms highly correlated with the presence of SARS-CoV-2 IgG, as well as the clinical definition of COVID-19 status (Table II). Overall, in univariate analyses (Table III), working in a COVID-19 ward (relative to working in a non-COVID-19 ward, 3.8% vs 3.2%, p = 0.50) or taking care of COVID-19 patients (4.0% vs 3.2%, p = 0.27), were not significantly associated with a positive status for SARS-CoV-2 IgG. Conversely, contact with a HCW who was diagnosed as COVID-19 was significantly associated with seropositivity for SARS-CoV-2 IgG (2.4% vs 4.9%, p=0.0004) as was household contact with someone diagnosed with COVID-19 (3.2% vs 6.9%, p = 0.0006). Activities associated with increased risk of positive IgG SARS-CoV-2 test in univariate analyses (Table IV) were clinical examination (p = 0.01), and the mobilisation of patients in bed (p = 0.01). Multivariate analyses (Table V), both using the ‘by combination’ or ‘by multiple imputation’ definitions, confirmed the association between SARS-CoV-2 IgG seropositivity and contact with HCWs diagnosed with COVID-19 (OR = 1.51 95%CI [1.18-1.94], by the ‘multiple imputation definition’) or relatives (OR = 1.42 95%CI [1.08-1.86]). On multivariate analyses adjusted for age, sex, occupation, and contact with a relative or patient with COVID-19, sub-optimal protective equipment during certain tasks was associated with positive test for SARS-CoV-2 IgG (‘by combination definition’): nasopharyngeal samplings (OR= 3.46 95%CI [1.15-10.40]), mobilisation of patient in bed (OR = 3.30 95%CI [1.51-7.25]), clinical examination (OR = 2.51 95%CI [1.16-5.43]) and eye examination (OR = 2.90 95%CI [1.01-8.35]). The same analyses using the ‘ by multiple imputation’ definition confirmed these associations only for mobilising patients and clinical examination (Table VI). Washing and dressing patients were also associated with increased risk of SARS-CoV-2 IgG seropositivity even with self-declared appropriate use of protective equipment, using both the ‘by combination’ and ‘by multiple imputation’ definitions (OR = 2.13 [95%CI 1.05-4.30] and OR= 1.51 95%CI [1.06-2.14], respectively). Finally, aerosol-generating procedures, whether with self-declared appropriate, or sub-optimal use of protective equipment were also associated with positive test for SARS-CoV-2 IgG using the ‘by multiple imputation’ method (OR=1.37 95%CI [1.04-1.81] and OR = 1.74 95%CI [1.05-2.88], respectively).

**Table I.**
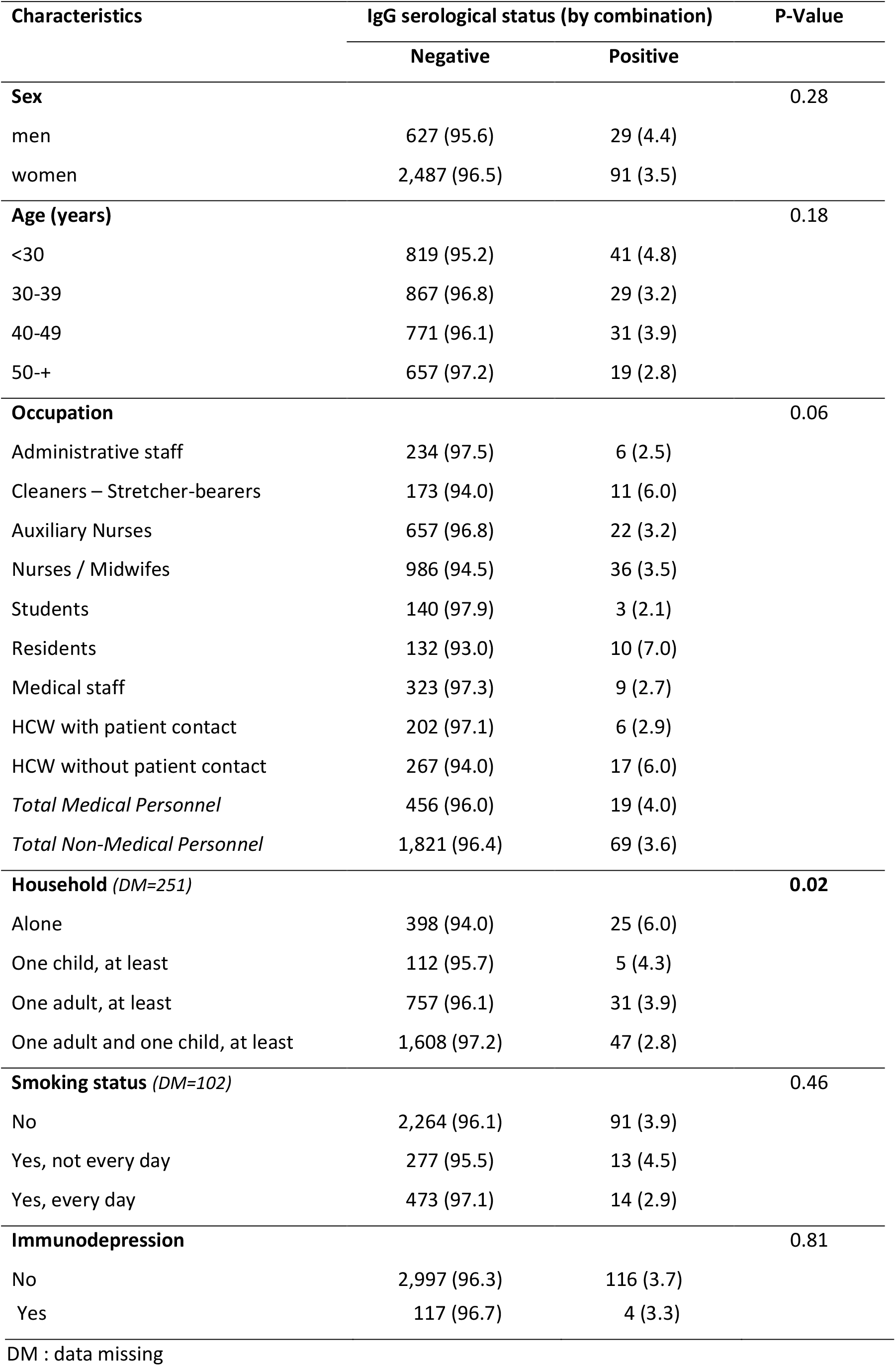
Overall characteristics of subjects enrolled in the HB AntiCov study (n=3,234)

**Table II.** Clinical characteristics of subjects enrolled in the HB AntiCoV study (N = 3,234)

| Characteristics | IgG serological status (by combination) |  | P-Value |
| --- | --- | --- | --- |
|  | Negative | Positive |  |
| **Symptoms** |  |  | **<0.0001** |
| No symptom | 1,103 (98.4) | 18 (1.8) |  |
| 1 | 459 (97.7) | 11 (2.3) |  |
| 2-3 | 826 (97.5) | 21 (2.5) |  |
| 4-6 | 553 (93.9) | 36 (6.1) |  |
| 7+ | 173 (83.6) | 34 (16.4) |  |
| Median | 1 | 4 | **<0.0001** |
| **COVID-19 (clinical status)** |  |  | **<0.0001** |
| No | 1,103 (98.4) | 18 (1.6) |  |
| Possible | 1,862 (98.0) | 39 (2.0) |  |
| Probable | 149 (70.3) | 63 (29.7) |  |
| **RT-PCR** |  |  | **<0.0001** |
| No test | 1,988 (98.2) | 36 (1.8) |  |
| Negative | 1,076 (97.6) | 26 (2.4) |  |
| Positive | 50 (46.3) | 58 (53.7) |  |
| **LFIAT (IgG) (DM=19)** |  |  | **<0.0001** |
| Negative | 3,026 (99.8) | 7 (0.2) |  |
| Positive | 69 (37.9) | 113 (62.1) |  |

298 DM : data missing, LFIAT: lateral flow immunoassay test

**Table III.**
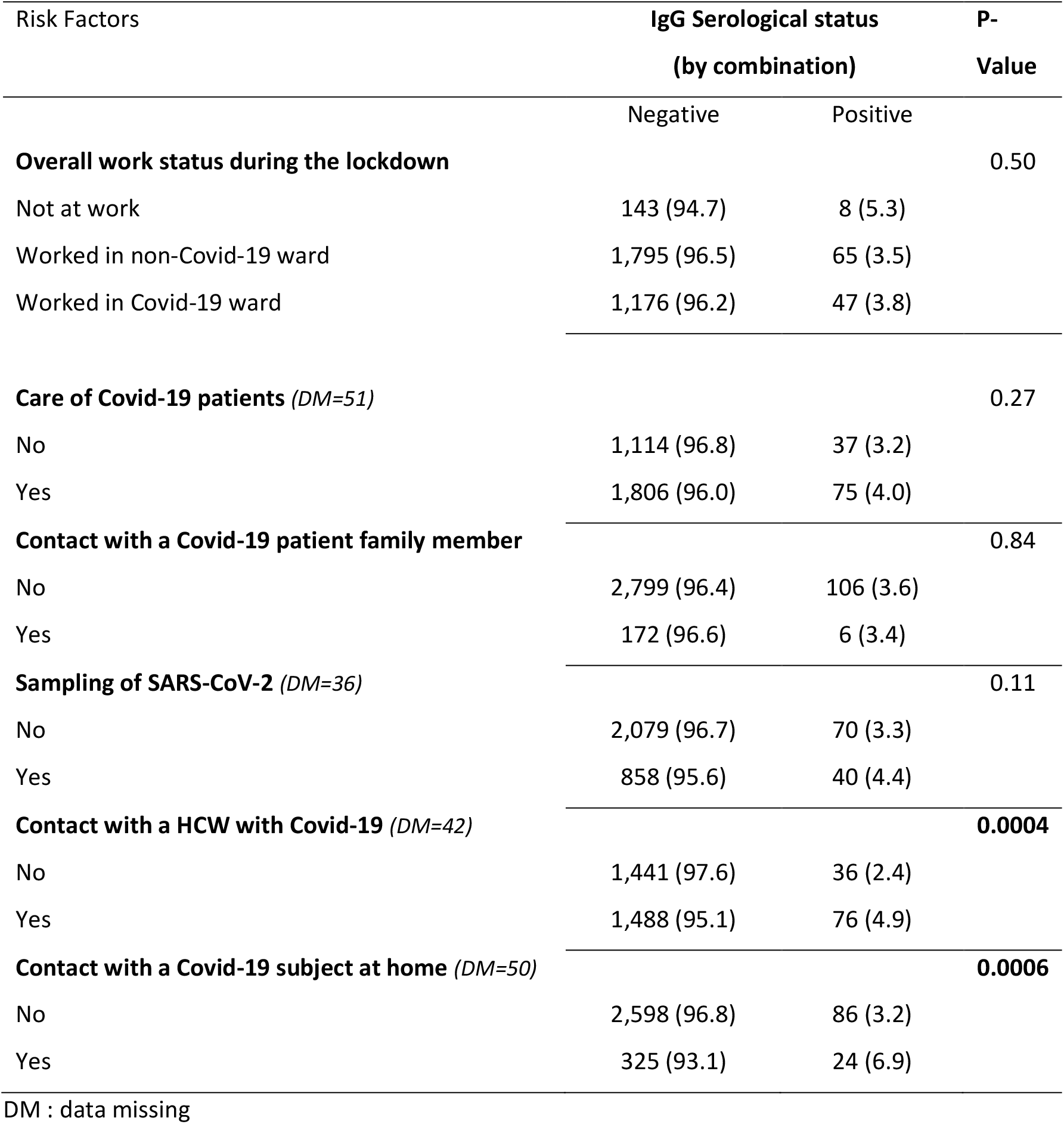
Overall risk Factors associated with SARS-CoV-2 IgG serological status obtained by combination in the HB AntiCoV Study (univariate analyses, n= 3,083)

**Table IV.**
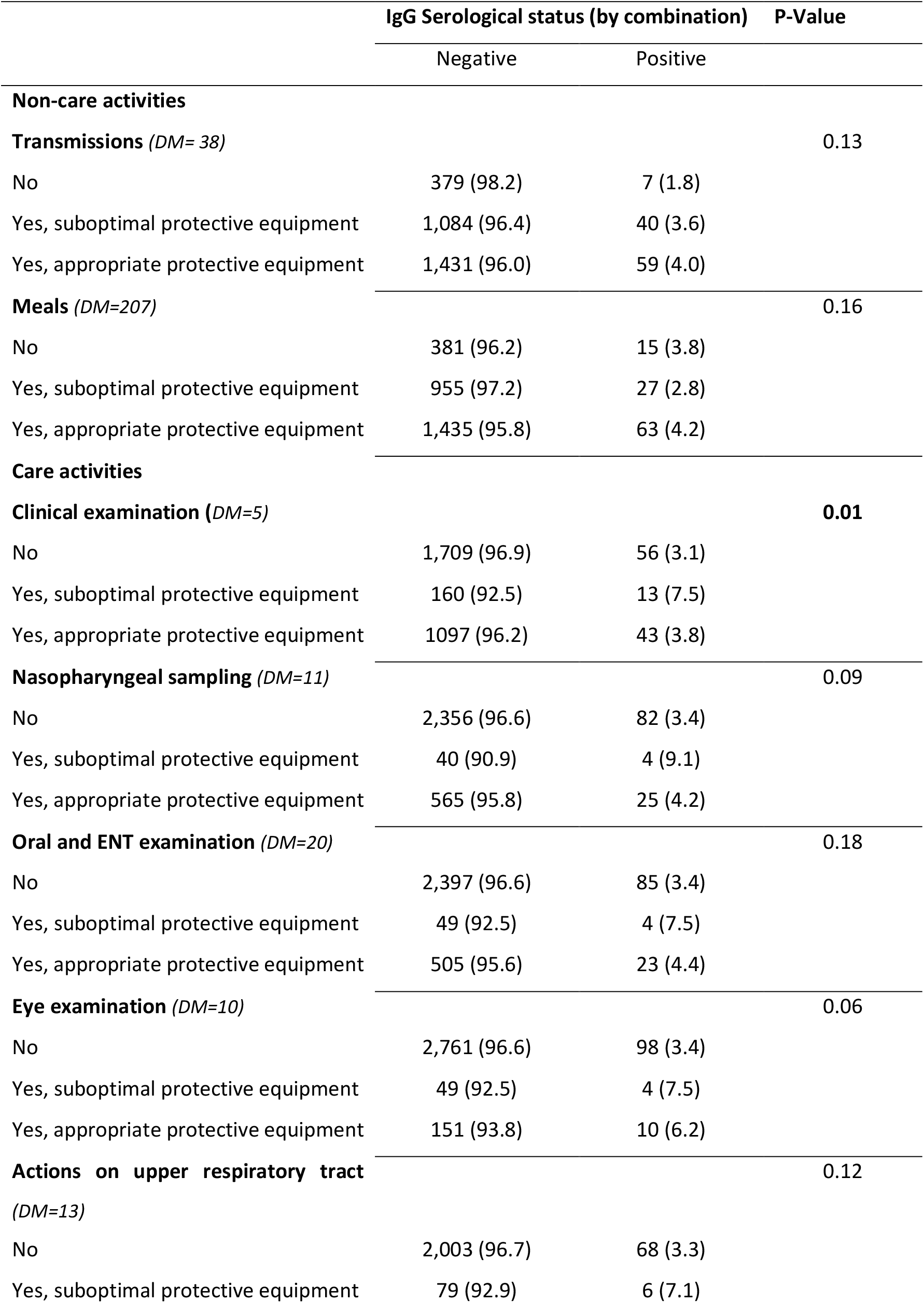
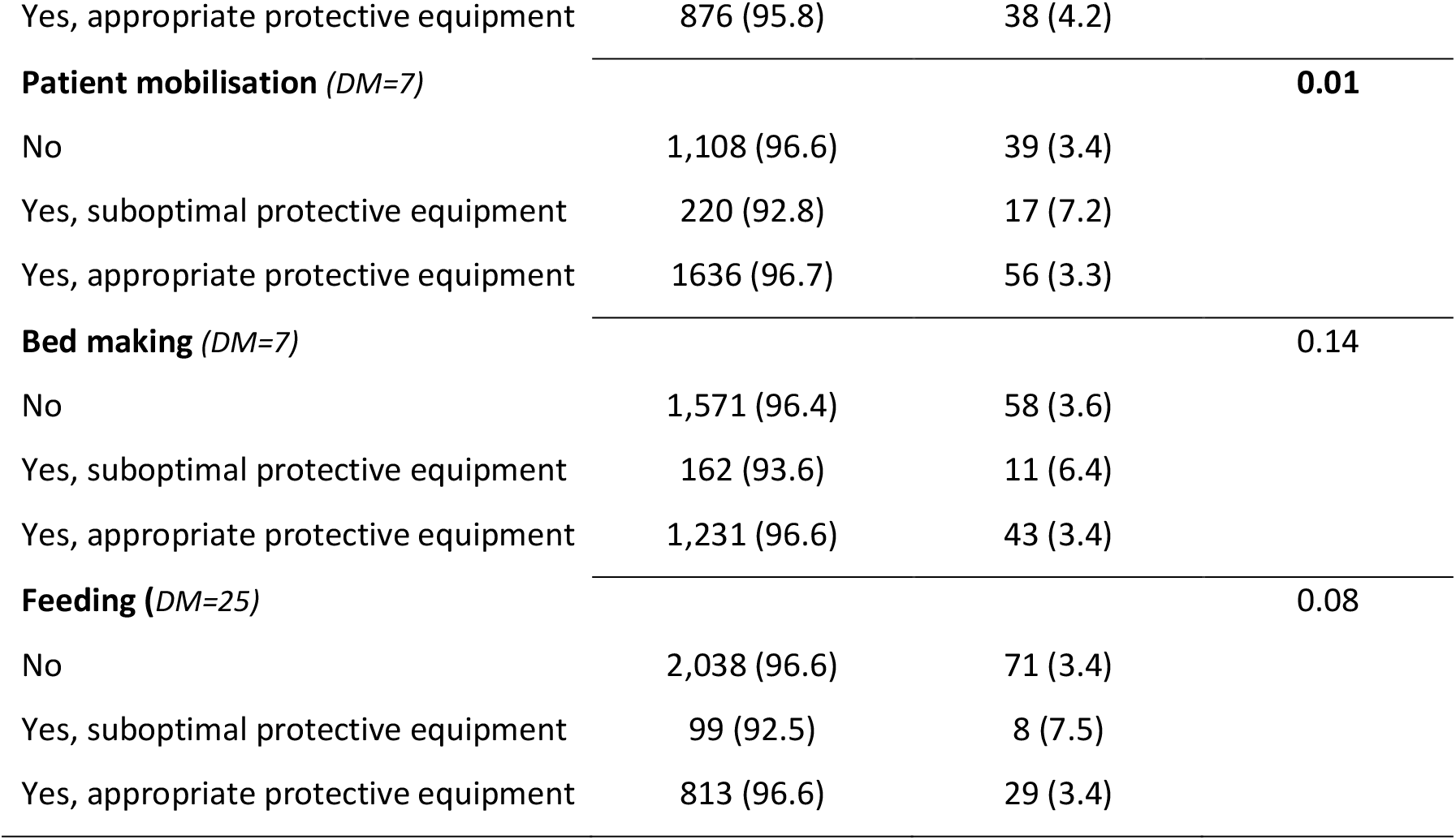
Specific tasks associated with the SARS-CoV-2 IgG serological status obtained by combination in the HB Anti-CoV Study (univariate analyses, n=3,083, P <0 .20)

**Table V.** Overall risk factors associated with the SARS-CoV-2 IgG serological status obtained by combination (n=2,866) or multiple imputation (n=50 data sets) in the HB Anti-CoV Study. *(Logistic regression models, adjusted for age, sex, occupation)*

| Variables | IgG serological status |  |
| --- | --- | --- |
|  | By combination | By multiple imputation |
|  | OR [CI 95%] | OR [CI 95%] |
| **Immunodepression (yes)** | 1.32 [0.47-3.74] | - |
| **Life at home (DM=251)** |  | ns |
| Alone | 1.00 |  |
| One child, at least | 0.54 [0.17-1.65] | - |
| One adult, at least | 0.58 [0.33-1.03] | - |
| One adult and one child, at least | **0.49 [0.28-0.85]** | - |
| **Care of Covid-19 patient (yes)** | 1.30 [0.78-2.17] | - |
| **Contact with family of a Covid-19 patient (yes)** | 0.90 [0.38-2.10] | - |
| **Covid-19 sampling (yes)** | 1.00 [0.63-1.60] | - |
| **Contact with a Covid-19 HCW (yes)** | **2.08 [1.33-3.24]** | **1.51 [1.18-1.94]** |
| **Contact with a Covid-19 subject at home (yes)** | **2.00 [1.23-3.28]** | **1.42 [1.08-1.86]** |

**Table VI.** Specific care tasks associated with the SARS-CoV-2 IgG serological status obtained by combination (n=2,961) or multiple imputation (n=50 data sets) in the HB Anti-CoV Study. (logistic regression, P-value <0.20, adjusted for age, sex, occupation, contact with a Covid-19 patient, contact with a Covid-19 subject at home) Each group of specific tasks by occupation corresponded to an independent model according to the factorial analysis (see methods)

| Variables | IgG Serological status |  |
| --- | --- | --- |
|  | by combination | by multiple imputation |
|  | OR [CI 95%] | OR [ICI 95%] |
| **Nurse specific tasks** |  |  |
| **Nasopharyngeal sampling** |  |  |
| No | 1.00 | 1.00* |
| Yes, suboptimal protective equipment | **3.46 [1.15-10.40]** | 0.88 [0.32-2.43] |
| Yes, appropriate protective equipment | 1.33 [0.79-2.25] | 1.13 [0.84-1.52] |
| **Central lines insertion** |  |  |
| No | **1.00** | **1.00** |
| Yes, suboptimal protective equipment | 2.66 [0.57-12.38] | 1.75 [0.82-3.74] |
| Yes, appropriate protective equipment | 0.72 [0.35-1.45] | 0.74 [0.48-1.14] |
| **Actions on upper respiratory tract** |  |  |
| No | 1.00* | 1.00 |
| Yes, suboptimal protective equipment | 1.85 [0.66-5.22] | **1.74 [1.05-2.88]** |
| Yes, appropriate protective equipment | 1.53 [0.92-2.55] | **1.37 [1.04-1.81]** |
| **Auxiliary Nurse specific tasks** |  |  |
| **Washing and dressing patient** |  |  |
| No | 1.00 | 1.00 |
| Yes, suboptimal protective equipment | 0.38 [0.10-1.46] | 0.63 [0.29-1.36] |
| Yes, appropriate protective equipment | **2.13 [1.05-4.30]** | **1.51 [1.06-2.14]** |
| **Patient mobilisation** |  |  |
| No | 1.00 |  |
| Yes, suboptimal protective equipment | **3.30 [1.51-7.25]** | **2.04 [1.33-3.11]** |
| Yes, appropriate protective equipment | 0.93 [0.49-1.79] | 1.00 [0.70-1.44] |
| **Feeding** |  |  |
| No | 1.00 | 1.00* |
| Yes, suboptimal protective equipment | 2.48 [0.92-6.72] | 1.24 [0.65-2.36] |
| Yes, appropriate protective equipment | 1.00 [0.54-1.89] | 0.87 [0.62-1.24] |
| **Specific Medical tasks** |  |  |
| **Clinical examination** |  |  |
| No | 1.00 |  |
| Yes, suboptimal protective equipment | **2.51 [1.16-5.43]** | **1.62 [1.06-2.48]** |
| Yes, appropriate protective equipment | 1.50 [0.85-2.64] | 1.23 [0.90-1.68] |
| **Eye Examination** |  |  |
| No | 1.00 | 1.00* |
| Yes, suboptimal protective equipment | **2.90 [1.01-8.35]** | 1.39 [0.66-2.91] |
| Yes, appropriate protective equipment | 1.86 [0.93-3.73] | 1.02 [0.60-1.72] |

\* : variable not included in the final regression logistic model as $p > 0.20$

## DISCUSSION

This large sero-epidemiological study, which included more than 3,000 HCWs in a French university hospital after the first epidemic wave, highlights several possible factors in the risk of transmission of SARS-CoV-2. First, our results confirm that, during the first epidemic wave, contact with both relatives or HCWs with COVID-19 were the two main risk factors for SARS-CoV-2 infection, while working in COVID-19 wards or contact with COVID-19 patients was not associated with an increased risk. Second, we confirmed that certain tasks performed by HCWs that increase the risk of aerosolization also increase the risk of SARS-CoV-2 transmission, particularly when use of protective equipment was sub-optimal, such as interventions on the upper respiratory tract or nasopharyngeal sampling. Our results also suggest that certain tasks associated with daily care, such as patients washing, dressing, mobilisation, and eye or clinical examinations also increased the risk of SARS-CoV-2 infection among HCWs.

A lower risk of transmission by working in COVID-19 wards during the first epidemic wave has already been reported. In a British cross-sectional study that included 545 HCWs, working in an intensive care unit with COVID-19 patiernts was associated with a lower risk of SARS-CoV-2 infection than working in other wards (OR = 0.28 95%CI [0.09-0.78] ^13^. Similarly, a large US cross-sectional study found no risk associated with working in COVID-19 wards (OR = 1.00 95%CI [0.98-1.03]) or intensive care units (OR = 0.98 95%CI [0.93-1.02]) among 49,329 HCWs tested for SARS-CoV-2 IgG antibodies ^14^. A study from China also reported a higher risk associated with working in non-COVID-19 wards (relative to dedicated wards, IRR = 3.1 95%CI [1.8-5.2]) ^15^. These findings suggest that appropriate use of protective equipment, better compliance and experience with COVID-19 in such dedicated wards would, indeed, be protective ^1, 16^. One study ^17^ reported an excess risk in frontline HCWs working in dedicated COVID-19 wards (RR= 1.65 95%CI [1.34-2.02]) among 28,792 subjects but that study considered both IgG and IgM antibodies obtained by a self-interpreted LFIA test to be positive, which may not be accurate. In a sero-prevalence study using a different LFIA test among 3,056 HCWs in a Belgian hospital, contact with COVID-19 patients was not associated with a higher prevalence of IgG antibodies against SARS-CoV-2 (OR = 1.08 95%CI [0.80-1.45] ^18^. In addition, Moscola et al. also did not observe a risk associated with direct patient care (RR = 95%CI [0.97-1.02]) ^14^. Conversely, Lentz et al. reported increased risk associated in HCWs with exposure to COVID-19 patients (OR=1.4 95%CI [1.0-1.9]), and that risk was associated with routine contact (1.4 95%CI [1.04-1.90]) rather than exposure to aerosol-generating procedures (OR =0.9 95%CI [0.6-1.2]) ^16^. Similarly, Shat et al. reported a higher risk of COVID-19 for HCWs facing patients (HR = 3.30 95%CI [2.13-5.13]) than those not facing patients, after adjustment for sex, age, ethnicity, socioeconomic status and comorbidity ^19^. Iversen et al. ^17^ also reported a mild excess risk for HCWs in contact with COVID-19 patients (RR = 1.22 95% CI [1.03-1.45]).

One explanation for such discrepancies may be the role of the appropriate use of protective equipment. Several at risk exposures have been reported for SARS, MERS, and, to a lesser extent, SARS-CoV-2. In a large literature review, Chou et al. highlighted that involvement in intubation, direct patient contact, and contact with bodily secretions increased the risk of coronavirus infections, but they found less convincing arguments for other types of exposure such as non-invasive positive-pressure ventilation, nebulizers use, manipulation of oxygen masks, and high-flow oxygen. This review confirmed the protective effect of using a mask, either surgical or N95^1^, previously reported ^20^. For coronaviruses, N95 masks are not generally found to be more protective than surgical masks for most at-risk exposures, albeit some authors reported better protection with N95 masks ^21, 22^. However, these studies suffered from methodological flaws. Our study suggests an increased risk associated with aerosol-generating procedures such as nasopharyngeal sampling in line with the recommendation to use N95 masks for these procedures. Moreover, actions on the upper respiratory tract were associated with a significant risk of SARS-CoV-2 infection, with or without protective equipment (OR = 1.37 95%CI [1.04-1.81] and OR = 1.74 95%CI [1.05-2.88], respectively using the ‘by multiple imputation’ definition). We also found a higher risk of SARS-CoV-2 IgG positivity associated with two auxiliary-nurses activities, namely patients washing and dressing, and their mobilisation. The masks used while performing these activities are surgical, and our results suggest that this level of protection may not be appropriate. One explanation may be that these activities require close, and prolonged contact with patients, two risk factors that may increase the risk of SARS-CoV-1 transmission ^21, 23, 24^. For SARS-CoV-2, to the best of our knowledge, only two studies suggested that duration of care may be a risk factor for infection. Thus, Lentz et al. documented an increased risk associated with care longer than 45 minutes ^16^. Grant et al., in their short paper, found a significant higher prevalence of SARS-CoV-2 antibodies among HCWs with exposure defined as prolonged direct contact with patients ^25^. Another explanation may be the generation of a small amount of SARS-CoV-2 aerosol ^26^ during such care activities, in particular by patients not wearing a mask, and contamination by inhalation, despite the use of a surgical mask. These findings suggest that protective equipment must be reinforced when HCWs are exposed to prolonged and close care of COVID-19 patients. Finally, we also observed a higher risk associated with eye and clinical examinations. These results are consistent with those of the literature ^7^, as SARS-CoV-2 can be detected in tears and conjunctival secretions ^27^ and eye examination requires close contact. Suboptimal use of protective equipment under these conditions may place HCWS at risk, as suggested by our results. This is the first study to suggest that clinical examination may be associated with increased risk even with self-declared appropriate use of protective equipment. This may also be possibly explained by close contact and, to a lesser extent, the duration of contact.

We also report increased risk of SARS-CoV-2 infection in HCWs with household relatives who were diagnosed with COVID-19 (OR = 1.42 95% CI [1.08-1.86]). Lentz et al. found a significant risk of infection associated with exposure outside of work, including living with a household member diagnosed with COVID-19 (OR = 3.8 CI95% [1.5-9.3]) or who presented COVID symptoms (OR = 3.1 CI95% [1.5-6.3]). Lai et al. also reported more frequent contact with confirmed cases of COVID-19 among family members than colleagues, albeit the difference was non-significant (12.7% *vs* 10.9%). Treibel et al. ^28^ compared the number and incidence of patients testing positive for SARS-CoV-2 in Greater London, to that of HCWs in their cohort study, and suggested that these data more likely reflect general community transmission than in-hospital exposure. Finally, Steensels et al. ^18^ found a significant association between sero-prevalence of SARS-CoV-2 antibodies and contact with suspected COVID-19 households (OR = 3.15 [95%CI 2.33-4.25]).

Our study has several limitations. First, data on the use of protective equipment, particularly masks, were only declarative, and some HCWs may have over- or under-reported their use. We tried to limit this effect by attributing the quality of protection independently of the tasks using the same algorithm throughout the database. However, we observed associations for only a few activities, and the observation of a coherent gradient of transmission risk with the quality of protection support the validity of our findings. Another limitation was the low sero-prevalence, resulting in a low statistical power. Nonetheless, we were able to highlight several activities associated with the risk of SARS-CoV-2 infection, even in multivariable analyses. We also only considered SARS-CoV-2 IgG as the LFIAT shows low performances for the detection of SARS-CoV-2 IgM ^11^. As we began our study at the end of May, two months after the peak of the first epidemic wave in France, the effect on our prevalence estimate was probably minimal. Finally, we only analysed the protective effect of masks, without considering gloves, visors, and lab coats. Thus, our results primarily focus mostly on the risk of of SARS-CoV-2 transmission by inhalation.

Our study also has several strengths. The determination of SARS-CoV-2 status was based on a LFIA test that we previously validated^12^. Our knowledge of the quality of both the negative and positive predictive values allowed us to include these data in our models, using multiple imputation after stratification on the LFIA test response. This method, generally used to complete missing data, was a good tool to correct our sero-prevalence results according to the validation measurement. Moreover, several authors have recommended accounting for such errors ^29, 30^. Thus, despite certain differences between the ‘in combination’ and ‘multiple imputation’ definitions, these approaches provide more confidence in our results.

Another strength was the selection of HCWs enrolled in the study. Our sample is representative of HCWs of our hospital as it included all HCWs within COVID-19 wards and a random selection of those working in non-COVID-19 wards. Moreover, the high rate of participation (80%) ensure that our sample was representative. A comparison of demographic and occupational characteristics between respondents and non-respondents did not find any difference (data not shown).

As previously mentioned, our study allowed us to precisely study the role of several care-associated activities, including nursing and auxiliary-nursing care. Our methodology, using factor analyses coupled to multivariate logistic regression also allowed us to take into account statistical correlations among the variables. However, residual correlations may explain some of the variation in the observed associations between specific care activities and the presence of SARS-COV-2 antibodies. As already mentioned and to the best of our knowledge, no previous study on this subject have used these statistical approaches.

## Conclusions

At a time during which second, and sometime third, wave of the SARS-CoV-2 pandemic is sweeping across the world, and during which many HCWs must care for COVID-19 patients, our study highlights several possible pathways of transmission associated with specific medical, nursing or auxiliary-nursing activities. Although effective protective equipment was already being used by HCWs, these findings support the possible role of less known situations in the transmission of SARS-CoV-2. In particular, long-duration, non-aerosol generating activities close to patients such as mobilising patients in their beds, washing and dressing them, and clinical or eye examinations may be at higher risk than previously thought. Better use of adequate protective equipment during these activities must be encouraged to better protect HCWs. Further studies are required to better understand the pathways of SARS-CoV-2 transmission among HCWs.

## Data Availability

Not applicable

## Contributors

CP and RG conceived the study. CP, RG, VT and PT contributed to the protocol and design of the study. ET, PGB, SO contributed to the implementation of the study or data collection. CH and VT conducted the ELISA serological assays. CP, AS, RG and PW conducted the statistical analysis. CP and RG contributed to the preparation of the report. All authors critically reviewed and approved the final version. All authors had full access to all the data in the study and had final responsibility for the decision to submit for publication.

## Acknowledgements

We thank all participants and nurses (Anne-Marie Le Bonniec, Sabine Filande, Veronique Grigoli, and Gaelle Oudard), and residents (Nadia Fatih, Alexandre Bichon, Jean Poinsignon, Annabelle Guilloux, Léah Rakotonirina and Ahmed Aiouaz) of the occupational medicine unit for inclusion of the participants. We thank Pr Boudjema, Pr Malledant, Nicolas Mevel, Sophie Huitorel, Agnes Gazzola, Anne-Sophie Jouault, and Valerie Turmel for their help in the organisation of this study.

