## Supplementary material for "Role of non-aerosols activities in the transmission of SARS-Cov-2 infection among health care workers": supll tables

Figure 1:

Flow-Chart

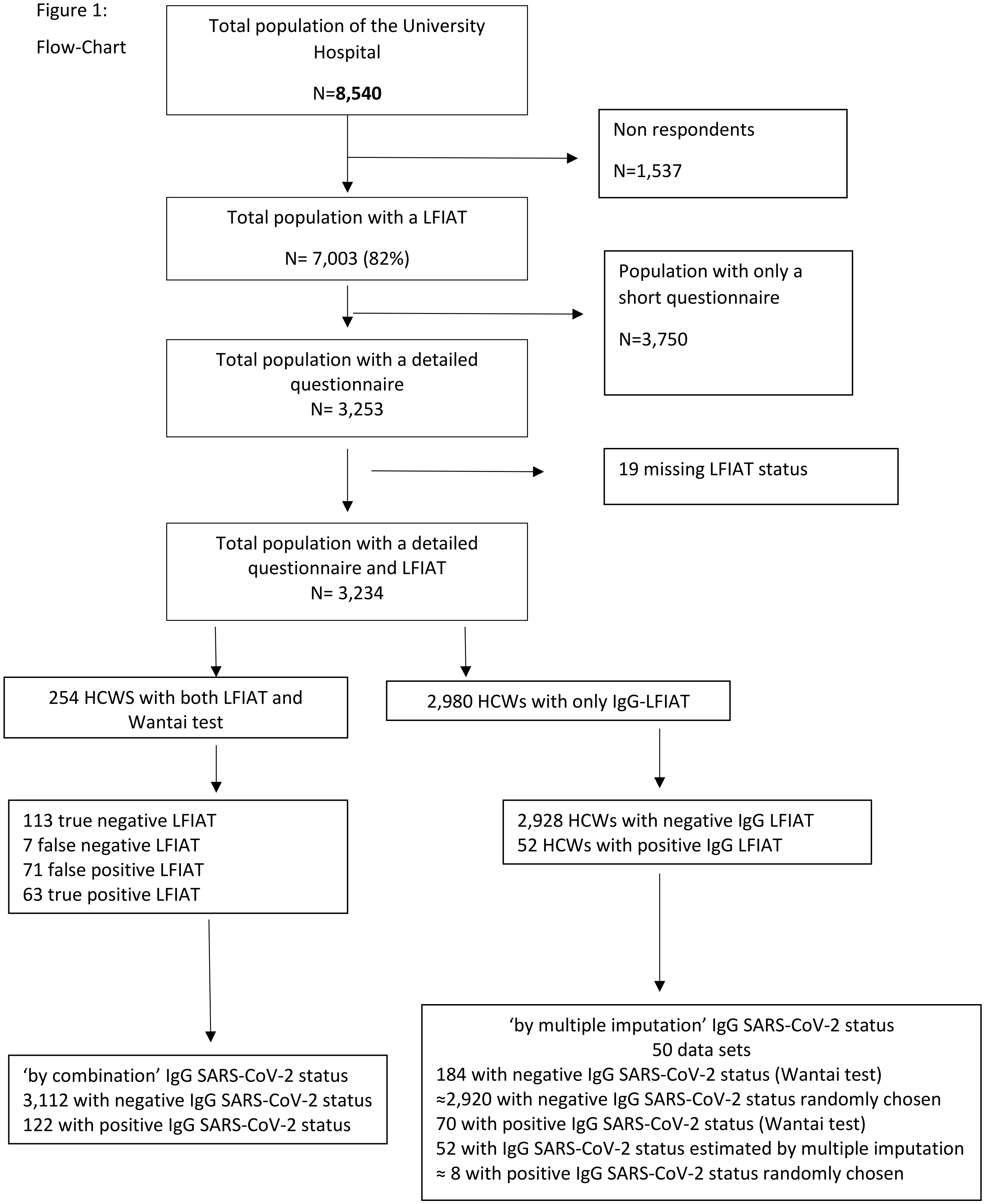

Suppl. Table I. Specific tasks associated with the SARS-CoV-2 IgG serological status obtained by combination in the HB Anti-CoV Study (univariate analyses, n=3,083, P >0 .20)

| | **IgG Serological status (by combination)** | | **P-Value** |
|---|---|---|---|
| **Non-care activities** | Negative | Positive | |
| **Changing room** *(DM=66)* | | | 0.50 |
| No | 847 (96.1) | 34 (3.9) | |
| Yes, suboptimal protective equipment | 939 (96.2) | 37 (3.8) | |
| Yes, appropriate protective equipment | 1125 (97.0) | 35 (3.0) | |
| **Meetings** *(DM=186)* | | | 0.43 |
| No | 1157 (96.9) | 37 (3.1) | |
| Yes, suboptimal protective equipment | 15 (93.8) | 1 (6.2) | |
| Yes, appropriate protective equipment | 1621 (96.1) | 66 (3.9) | |
| **Breaks** *(DM=429)* | | | 0.24 |
| No | 733 (96.5) | 27 (3.5) | |
| Yes, suboptimal protective equipment | 292 (98.3) | 5 (1.7) | |
| Yes, appropriate protective equipment | 1539 (96.4) | 58 (3.6) | |
| **Care activities** | | | |
| **Consulting** *(DM=1)* | | | 0.39 |
| No | 475 (96.2) | 19 (3.8) | |
| Yes, suboptimal protective equipment | 288 (95.1) | 15 (4.9)) | |
| Yes, appropriate protective equipment | 2207(96.6) | 78 (3.4) | |
| **Vitals (***DM=4)* | | | 0.95 |
| No | 1433 (96.4) | 53 (3.6) | |
| Yes, suboptimal protective equipment | 172 (96.6) | 6 (3.4) | |
| Yes, appropriate protective equipment | 1362 (96.3) | 53 (3.7) | |
| **Central line insertion** *(DM=24)* | | | 0.22 |
| No | 2587 (96.3) | 100 (3.7) | |
| Yes, suboptimal protective equipment | 18 (90.0) | 2 (10.0) | |
| Yes, appropriate protective equipment | 342 (97.2) | 10 (2.8) | |
| **Peripheral IV insertion** *(DM=16)* | | | 0.93 |

| | | | |
|---|---|---|---|
| No | 1936 (96.3) | 74 (3.7) | |
| Yes, suboptimal protective equipment | 114 (95.8) | 5 (4.2) | |
| Yes, appropriate protective equipment | 905 (96.5) | 33 (3.5) | |
| **Urinary catheter insertion (***DM=14)* | | | 0.47 |
| No | 2328 (96.6) | 83 (3.4) | |
| Yes, suboptimal protective equipment | 54 (94.7) | 3 (5.3) | |
| Yes, appropriate protective equipment | 575 (95.7) | 26 (4.3) | |
| **Nasogastric tube insertion** *(DM=10)* | | | 0.33 |
| No | 2435 (96.3) | 95 (3.7) | |
| Yes, suboptimal protective equipment | 55 (100.0) | 0 (0.0) | |
| Yes, appropriate protective equipment | 471 (9657) | 17 (3.5) | |
| **Delivery** *(DM=13)* | | | 0.44 |
| No | 2836 (96.3) | 110 (3.7) | |
| Yes, suboptimal protective equipment | 14 (100.0) | 0 (0.0) | |
| Yes, appropriate protective equipment | 108 (98.2) | 2 (1.8) | |
| **Loco-regional anaesthesia** *(DM=24)* | | | 0.31 |
| No | 2846 (96.4) | 107 (3.6) | |
| Yes, suboptimal protective equipment | 18 (90.0) | 2 (10.0) | |
| Yes, appropriate protective equipment | 83 (96.5) | 2 (3.5) | |
| **Surgery** *(DM=12)* | | | 0.35 |
| No | 2726 (96.2) | 107 (3.8) | |
| Yes, suboptimal protective equipment | 26 (100.0) | 0 (0.0) | |
| Yes, appropriate protective equipment | 207 (97.6) | 5 (2.4) | |
| **Distribution of drugs** *(DM=12)* | | | 0.80 |
| No | 2019 (96.4) | 76 (3.6) | |
| Yes, suboptimal protective equipment | 142 (97.3) | 4 (2.7) | |
| Yes, appropriate protective equipment | 798 (96.1) | 32 (3.9) | |
| **Washing and dressing patient** *(DM=13)* | | | 0.63 |
| No | 1814 (96.6) | 64 (3.4) | |
| Yes, suboptimal protective equipment | 111 (96.5) | 4 (3.5) | |
| Yes, appropriate protective equipment | 1033 (96.6) | 44 (4.1) | |
| **Mouth washing** *(DM=17)* | | | 0.40 |
| No | 2073 (96.9) | 72 (3.4) | |

| | | | |
|---|---|---|---|
| Yes, suboptimal protective equipment | 84 (95.5) | 4 (4.5) | |
| Yes, appropriate protective equipment | 797 (95.7) | 36 (4.3) | |
| **Physiotherapy (mobilisation)** *(DM=29)* | | | 0.90 |
| No | 2847 (96.4) | 106 (3.6) | |
| Yes, suboptimal protective equipment | 24 (96.0) | 1 (4.0) | |
| Yes, appropriate protective equipment | 74 (97.4) | 2 (2.6) | |
| **Physiotherapy (respiratory) (***DM=32)* | | | 0.97 |
| No | 2880 (96.5) | 106 (3.5) | |
| Yes, suboptimal protective equipment | 62 (96.9) | 2 (3.1) | |
| Yes, appropriate protective equipment | 1 (100.0) | 0 (0.0) | |
| **Dental treatment (***DM= 32)* | | | 0.92 |
| No | 2810 (96.5) | 103 (3.5) | |
| Yes, suboptimal protective equipment | 46 (95.8) | 2 (4.2) | |
| Yes, appropriate protective equipment | 92 (95.8) | 4 (4.2) | |
